# Pathogenic PF4/Polyvinylsulfonate ELISA-negative Antibodies in HIT

**DOI:** 10.1101/2025.10.29.25338884

**Authors:** Adam J. Kanack, Noah P. Splinter, Emily E. Mauch, Leben Tefera, Morayma Reyes Gil, Sakshi Jasra, Andrew Goodwin, Kristi J Smock, Hadiyah Ahmad, Aneel Ashrani, Nathaniel L. Robinson, Ana I. Casanegra, Curtis G. Jones, Shannon M. Pechauer, Erin Yttre, Richard H. Aster, Mindy C. Kohlhagen, Rachel R. Leger, David L. Murray, Lu Zhou, Demin Wang, Renren Wen, Dong Chen, Rajiv K. Pruthi, Anand Padmanabhan

## Abstract

**BACKGROUND:** Platelet factor 4-polyanion enzyme-linked immunosorbent assays (ELISAs) are considered highly sensitive for diagnosing heparin-induced thrombocytopenia (HIT), such that current practice guidelines recommend use of ELISA-negative results to exclude HIT. Once HIT is ruled out, alternative, non-heparin-based anticoagulant treatments are ceased, and heparin reintroduction frequently occurs.

**METHODS:** Antigen-based and PF4-dependent functional testing were used to study PF4/polyvinyl sulfonate ELISA-negative platelet-activating antibodies in HIT-suspected patients and mice immunized with PF4/heparin.

**RESULTS:** Three patients with clinical presentations consistent with HIT tested negative in an ELISA using PF4-polyvinylsulfonate (PF4/PVS), an antigenic target very commonly used for HIT antibody detection. All three patients demonstrated PF4-dependent platelet activation in functional testing that was sensitive to blockade of platelet FcγRIIa receptors and inhibited by high concentrations of heparin, consistent with pathogenic HIT antibodies. Functional testing-based screening of 500 ELISA-negative patients identified three patients whose sera activated platelets in a PF4- and FcγRIIa-dependent manner, and had clinical histories consistent with HIT. Five of the six ELISA-negative HIT patients were re-exposed to heparin, which precipitated a decrease in platelet counts in all re-exposed patients, and one patient developed a new thrombus. To advance the study of ELISA-negative HIT antibodies, mice were immunized with PF4/heparin, and functional and antigenic assays were simultaneously used to successfully identify an ELISA-negative, PF4-dependent platelet-activating murine monoclonal antibody that recapitulated the serological characteristics of ELISA-negative HIT patients.

**CONCLUSIONS:** Recognition of ELISA-negative HIT is critical to avoid harm due to the cessation of alternative anticoagulation therapy and re-exposure of these patients to heparin.

---

Antibodies to Platelet factor 4 (PF4) are implicated in heparin-induced thrombocytopenia (HIT)^1–3^, and several recently identified syndromes, including vaccine-induced immune thrombotic thrombocytopenia (VITT)^4–9^ and monoclonal gammopathy of thrombotic significance (MGTS)^10^. In disorders of this family, including the prototypical disorder, HIT, frontline enzyme-linked immunosorbent assays (ELISAs) are used to evaluate antibody reactivity to PF4-polyanion complexes. Based on the high negative predictive value of these assays for detecting HIT antibodies^2^, guidelines from professional medical associations such as the American Society of Hematology recommend cessation of non-heparin anticoagulation therapy and the resumption of heparin (if indicated) in patients suspected of HIT with negative ELISA results (Recommendation 2.7), including those with high 4Ts scores (Recommendation 2.8)^11^. We encountered three index patients with ELISA-negative platelet-activating anti-PF4 antibodies, with clinical courses highly consistent with HIT. Given these findings, this study sought to evaluate the impact of heparin reintroduction that occurred in these ELISA-negative HIT patients and examine the incidence of ELISA-negative HIT in a large cohort of consecutive HIT-suspected patients who tested negative in one of the most frequently used HIT ELISAs with PF4-polyvinylsulfonate antigenic targets^12^.

## METHODS

### Patient Samples

Blood samples from index patients suspected of HIT and 500 consecutive samples from patients suspected of HIT at Mayo Clinic who tested negative in the Lifecodes PF4 IgG assay (Immucor) were used in research testing. The studies were approved by Mayo Clinic’s Institutional Review Board.

### ELISA and platelet studies

Lifecodes PF4 IgG and PF4 GAM (Immucor), Asserachrom HPIA GAM (Diagnostica Stago), and Zymutest HIA IgG (Hyphen BioMed) immunoassays were performed according to manufacturer instructions. The PF4-dependent P-selectin expression assay (PEA), a validated technique currently used as a diagnostic test for HIT in the US, and studies to evaluate IgG binding to platelets were performed as previously described^13–15^ and as detailed in the *Supplementary Appendix*. Serotonin release assay (SRA) testing was conducted at various reference laboratories, as ordered by the treating physician.

### ELISA-negative HIT murine monoclonal antibody development

Mice were immunized with PF4/unfractionated heparin, and sera/hybridomas were screened in the PF4-dependent P-selectin expression assay (PEA) and PF4/polyvinylsulfonate ELISA as detailed in the *Supplementary Appendix*.

## RESULTS

The first index case of ELISA-negative HIT (EN-HIT1; **Fig. 1A**) was a male in his 70s who was started on unfractionated heparin (UFH) for ventricular tachycardia and multivessel coronary artery disease. As shown in **Fig. 1A**, the patient developed thrombocytopenia with a 4Ts score of 6 and thrombosis of the left iliac and femoral arteries. Heparin anticoagulation was switched to a direct thrombin inhibitor (DTI) while ELISA testing was pending. Upon receipt of a negative ELISA [optical density (OD), 0.260; positive cut-off: ≥0.4 OD], unfractionated heparin treatment was re-commenced and was complicated by the development of new aortic and left popliteal artery thrombosis (**Fig. 1A**). Repeat ELISA testing was negative (OD, 0.175). SRA testing yielded positive results for one sample (44%, post-heparin day 11; positive cut-off ≥20%) and negative results for a second sample (7%, post-heparin day 15). The patient was transitioned to a DTI upon receipt of the weak-positive SRA results. After a complicated course, the patient required a left above-knee amputation.

**Figure 1.**
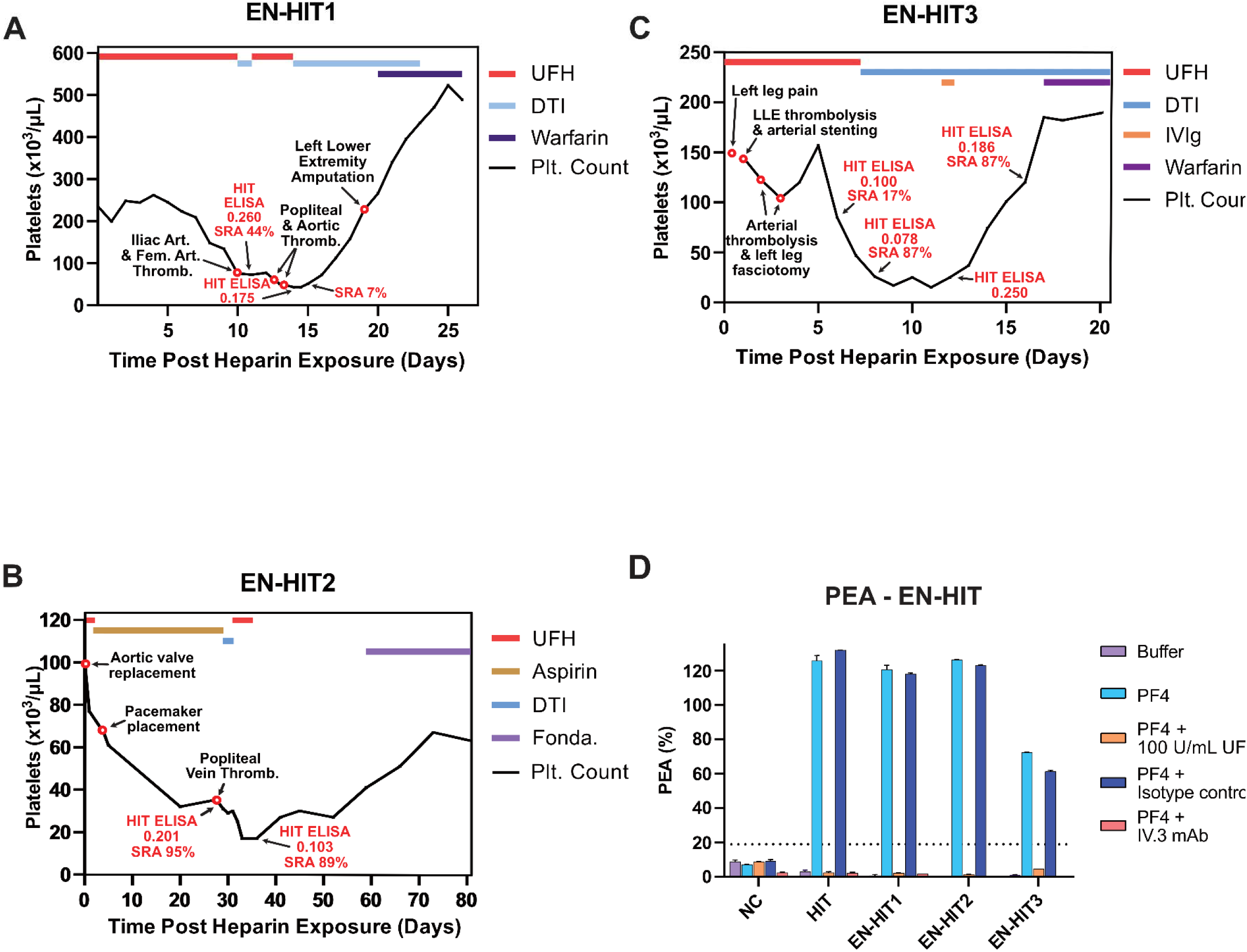
Clinical course of the three index ELISA-negative HIT patients. The clinical courses of ELISA-negative (EN)-HIT patients 1-3 are presented in **(A-C)**. The elapsed time post-heparin exposure is displayed on the x-axis, and platelet counts are displayed on the y-axis of each panel. Red circles denote major clinical events, and diagnostic testing results are in red font. The timeline of administration for the medications used to treat each patient is listed at the top of each graph and is denoted in the legend of each panel. Treatment with a direct thrombin inhibitor is denoted by DTI, and treatment with intravenous immunoglobulin G by IVIg. **(D)** PF4-dependent P-selectin expression assay (PEA) testing results are displayed for each of the three index EN-HIT patients in several testing conditions: buffer-treated platelets, PF4-treated platelets, PF4-treated platelets with excess (100 U/mL) unfractionated heparin (UFH), PF4-treated platelets with an isotype control (IC) monoclonal antibody (2 µg/mL), or PF4-treated platelets with the FcγRIIa-blocking monoclonal antibody IV.3 (2 µg/mL). NC-normal serum control; HIT-HIT positive control. NC-normal serum control; HIT-HIT positive control. Displayed PEA results **(D)** are the means of triplicate measurements with error bars representing one standard deviation.

The second index case, EN-HIT2 (**Fig. 1B**), was a male in his 60s with a history of mild thrombocytopenia possibly secondary to short telomere syndrome and prior lung transplantation for idiopathic pulmonary fibrosis who underwent a transcatheter aortic valve replacement. During this procedure, he received unfractionated heparin. In the days following surgery, he experienced a gradual decline in his platelet counts. One month after the procedure, the patient was noted to have worsening thrombocytopenia (35 x 10^3^/µL) and elevated D-dimer levels (1084 ng/mL FEU). Doppler ultrasound testing demonstrated a thrombus in the right popliteal vein. Due to high suspicion of HIT (4Ts score of 6), a DTI was initiated. However, HIT ELISA testing returned negative (OD, 0.201), prompting the resumption of heparin, which was followed by worsening thrombocytopenia (**Fig. 1B**). SRA results were strongly positive (95%), and repeat HIT testing showed a similar pattern (ELISA-negative but SRA-positive). Due to the significant thrombocytopenia and concern for bleeding, DTI treatment was stopped until platelet counts underwent a partial recovery, after which the patient was treated with fondaparinux (**Fig. 1B)**.

The third case, EN-HIT3 (**Fig. 1C**), was a male in his 50s with a history of left common iliac artery angioplasty and stent placement presenting with increasing leg pain, numbness, and changes in skin temperature. He had a baseline platelet count of 145 x10^3^/µL and was treated with UFH and balloon angioplasty/tissue plasminogen activator for stent and tibial artery occlusions. He had a steady decline in platelet count, reaching a nadir of 15 x10^3^/µL 11 days after heparin treatment, associated with re-occlusion of previously thrombolyzed vessels (**Fig. 1C**) and complicated by compartment syndrome, which was treated by fasciotomy. HIT ELISA testing was consistently negative (x3) with SRA-positive results at two of the three timepoints tested (**Fig. 1C**). The 4Ts score was 7. Platelet count recovery occurred only after treatment with intravenous immunoglobulin G (IVIg), suggesting the presence of a highly pathogenic HIT antibody.

All three patients demonstrated PF4-dependent platelet activation that was inhibited by a high concentration of heparin, typical of anti-PF4 antibodies (**Fig. 1D**). Additionally, the murine monoclonal antibody (mab) IV.3, which blocks the interaction of anti-PF4 antibodies with the platelet IgG receptor, FcγRIIa, effectively inhibited activation induced by all three patient samples, confirming the presence of platelet-activating IgG antibodies (**Fig. 1D**).

As a logical next step, we sought to define the incidence of pathogenic platelet-activating ELISA-negative anti-PF4 antibodies among a large cohort of consecutive HIT-suspected patients with negative PF4/PVS ELISA results. In this initial screen using the PF4-dependent p-selectin expression assay (PEA), 20 of the 500 patient samples stimulated platelet activation (PEA ≥19%; **Fig. 2A**). To exclude non-HIT antibodies (e.g., HLA antibodies) as the cause of activation, all 20 of the initial positives were evaluated for PF4-dependence and inhibitability with high concentrations of heparin, which are hallmarks of pathogenic anti-PF4 antibodies. The results of this secondary screen demonstrated that 17 out of 20 patient samples did not have a HIT-like profile (in fact, several did not reproducibly activate platelets obtained from a different donor, suggesting other platelet-directed antibodies (e.g., anti-HLA) as the possible cause of positivity in the primary screen; **Supplementary Fig. S1**). Samples from three PF4/PVS ELISA-negative patients (EN-HIT4-6), demonstrated a classical PF4-dependent, mab IV.3 and high concentration heparin inhibited profile, consistent with pathogenic platelet-activating HIT antibodies (**Fig. 2B**). Upon retrospective review, all three patients identified by the screen had clinical courses consistent with HIT (see *Supplemental Appendix* for detailed histories). Due to negative ELISA results, all three patients were re-exposed to heparin, and all patients exhibited a robust decrease in platelet count upon reintroduction, further supporting a pathogenic anti-PF4 antibody-mediated HIT process (**Fig. 2C-E**). The SRA was performed retrospectively and was negative (<5%) in EN-HIT4, positive (90%) in EN-HIT5, and not performed due to inadequate sample volume in EN-HIT6.

**Figure 2.**
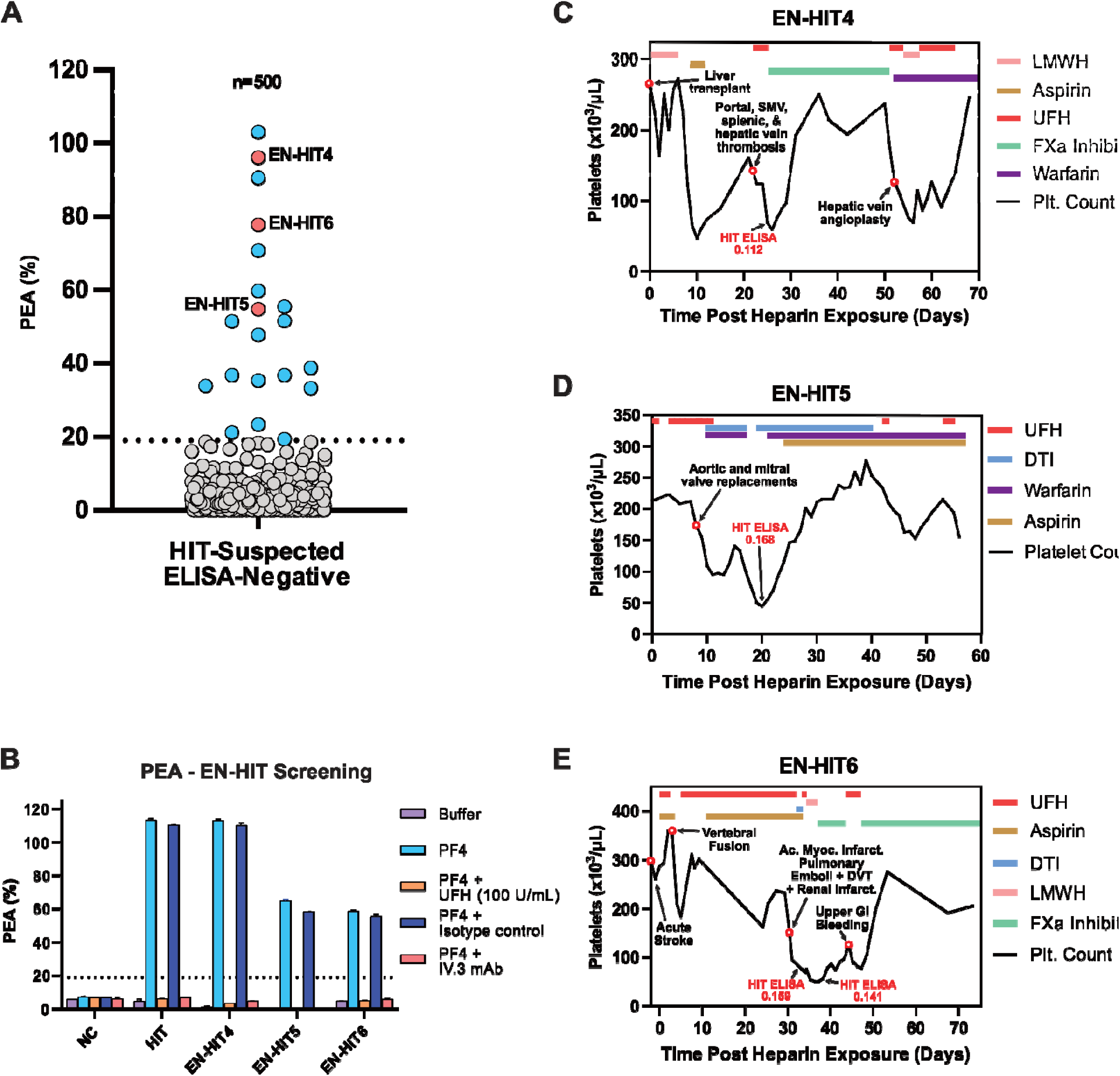
Three patients with pathogenic anti-PF4 antibodies were identified from a screen of 500 consecutive HIT-suspected patients who were ELISA-negative. **(A)** An initial PF4-dependent P-selectin expression assay (PEA) screen of 500 consecutive ELISA-negative (Lifecodes PF4 IgG, Immucor) patients yielded 20 patient samples (blue and orange circles) that stimulated platelet activation (≥19%). **(B)** Confirmatory PEA testing was performed using platelets from a second, independent platelet donor using several conditions: buffer-treated platelets, PF4-treated platelets, PF4-treated platelets with excess (100 U/mL) unfractionated heparin (UFH), PF4-treated platelets with an isotype control (IC) monoclonal antibody (2 µg/mL), or PF4-treated platelets with the FcγRIIa-blocking monoclonal antibody IV.3 (2 µg/mL). The three EN-HIT patients that demonstrated IgG-mediated PF4-dependent platelet activation, inhibitable with high concentrations of heparin, are denoted in **(A)** by orange circles. The clinical courses of these three additional patients (EN-HIT patients 4-6) are presented in **(C-E)**. The elapsed time post-heparin exposure is displayed on the x-axis, and platelet counts are displayed on the y-axis of each panel. Red circles denote major clinical events, and diagnostic testing results are in red font. The treatment of patients with medications is displayed at the top of each graph and is shown in the legend of each panel. LMWH, low molecular weight heparin; DTI, direct thrombin inhibitor; DVT, deep vein thrombosis.

Testing demonstrated PF4-dependent binding of five of the six patient antibodies to platelets, as has been demonstrated previously for ELISA-positive HIT antibodies (one sample, EN-HIT6, was not tested due to inadequate volume)^15^ (**Fig. 3A**). PF4-dependent platelet binding was also inhibited by high concentrations of heparin, consistent with the platelet activation profiles presented in **Figures 1 and 2**. All antibodies were negative in an IgG-specific PF4-polyvinylsulfonate target-based ELISA (Lifecodes PF4 IgG), as described in the patient histories. In the polyspecific version of this test, which detects IgG, IgA and IgM HIT isotypes, two of the six samples were weakly positive (Optical density, 0.722 and 0.523, Lifecodes PF4 IgGAM, **Fig. 3B and Table 1**). The use of an FDA-approved ELISA with PF4-heparin targets (Asserachrom HPIA IgGAM) also resulted in the reliable detection of only two of the six patient antibodies. Two patients, EN-HIT1 and EN-HIT3, produced results very close to (below and above) the cut-off in two independent runs, and were deemed negative based on the calculated mean ODs and cut-offs of two separate runs [mean 0.34 (cut-off, 0.38), and 0.32 (cut-off, 0.38); **Fig. 3C and Table 1**]. Finally, a fourth FDA-approved diagnostic assay using platelet lysate-heparin targets led to the identification of five of the 6 antibodies (Zymutest IgG, **Fig. 3D, and Table 1**). Samples with adequate available volumes were also tested in a recently described “off-the-shelf” PF4-dependent cryopreserved platelet assay that measures Thrombospondin-1 (TSP-1) release as a marker of platelet activation^16^ (**Fig. 3E**). This assay detected antibodies in both samples tested (EN-HIT4 and EN-HIT5), consistent with PEA results. **Table 1** summarizes HIT serological testing and 4Ts scores of the six patients in our study. To assess the presence of antibodies to non-PF4 targets, such as IL-8 and NAP-2, which have been previously suggested as potential alternative targets for HIT antibodies, samples available in adequate volumes (EN-HIT 1-5) were tested in ELISAs to assess binding to NAP-2 and IL-8. Results demonstrate that only one patient (EN-HIT3) appeared to have antibodies to IL-8, but the optical density was not higher than those observed with some ELISA-positive HIT patients, bringing into question the significance of this finding (**Supplementary Fig. S2**).

**Figure 3.**
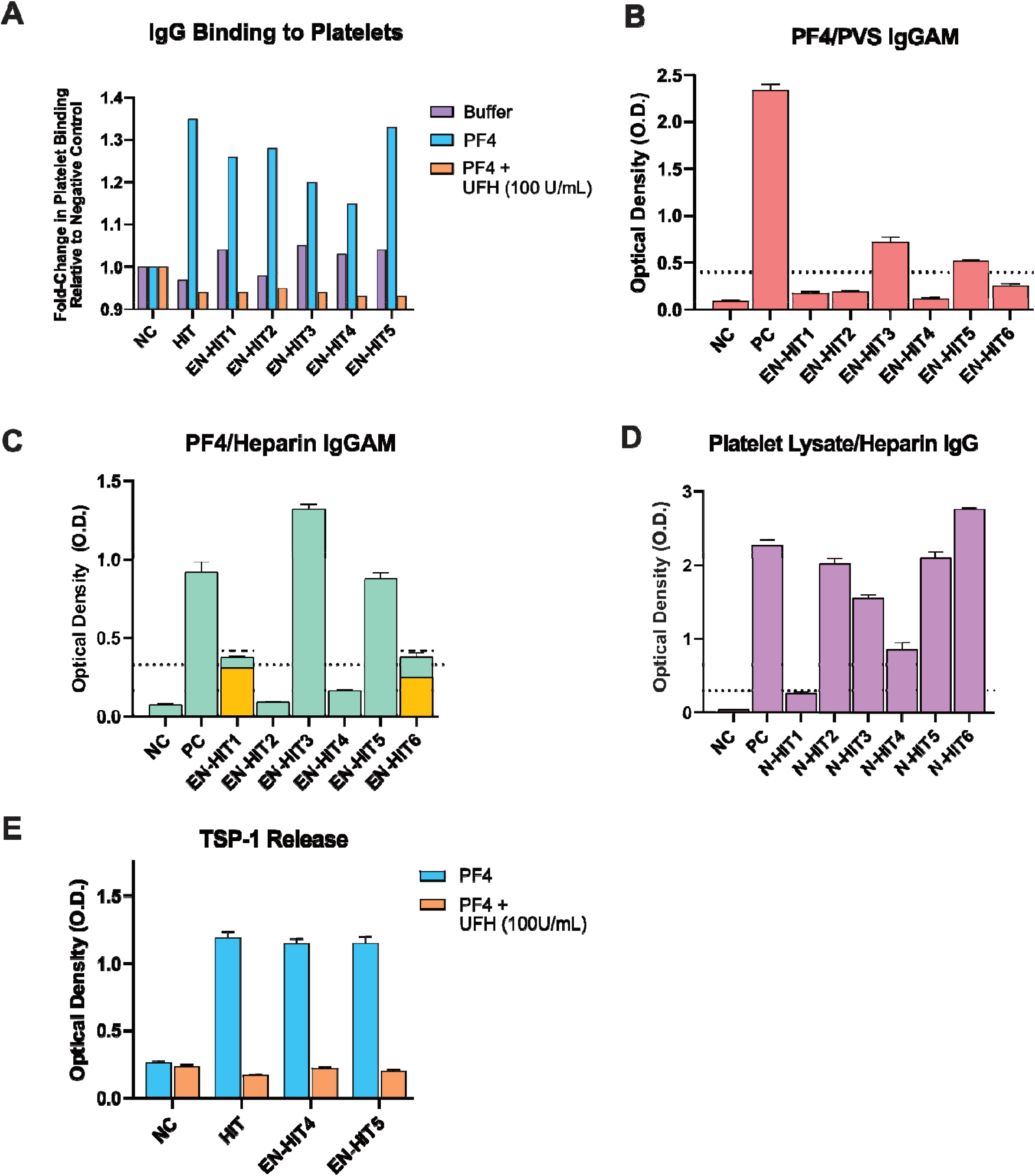
Serologic characterization of ELISA-negative HIT patients. **(A)** IgG binding to platelets was quantified after incubating PF4-treated platelets with sera from EN-HIT patients. Platelets were incubated with buffer alone, PF4-treated platelets, or PF4-treated platelets with 100 U/mL unfractionated heparin (UFH). Due to volume limitations, each platelet binding condition was only performed in singleton. NC, normal control; HIT/PC, positive control. HIT ELISAs were performed according to manufacturer instructions: **(B)** PF4/PVS polyspecific ELISA-Lifecodes PF4 Ig GAM (Immucor), **(C)** PF4/Heparin polyspecific ELISA-Asserachrom HPIA GAM (Diagnostica Stago). Two patients, EN-HIT1 and EN-HIT6, each tested borderline positive and negative in two independent test runs of the Asserachrom HPIA GAM and were deemed negative due to low optical densities, and **(D)** Platelet lysate/heparin IgG-specific ELISA-Zymutest HIA IgG immunoassay (Hyphen BioMed). Manufacturer-defined positive cut-offs are displayed for each assay by a dashed line. Each bar represents the mean, and error bars represent one standard deviation. **(E)** Detection of Thrombospondin-1 (TSP-1) release induced by EN-HIT4 and ENHIT5 samples using PF4-treated “off-the-shelf” cryopreserved platelets^16^ was evaluated in ELISA format. Each bar represents the mean, and error bars represent one standard deviation.

**Table 1.**
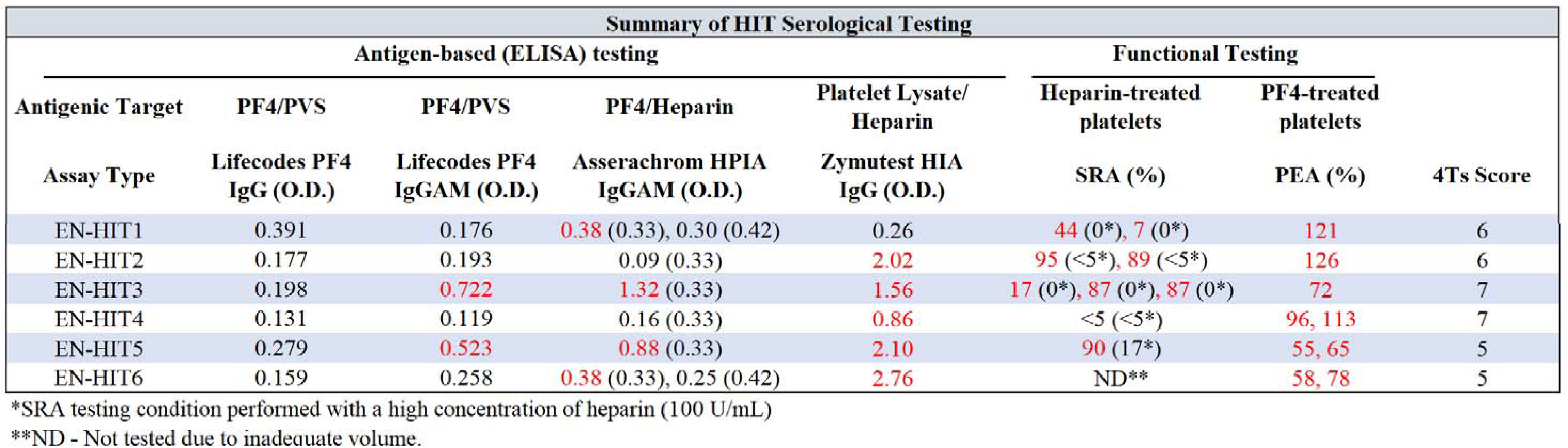
Summary of anti-PF4 serologic testing in six PF4/PVS ELISA-negative (EN)-HIT patients. Results from several HIT immunoassays [PF4/PVS-Lifecodes PF4 IgG and PF4 IgGAM (Immucor), PF4-Heparin-Asserachrom HPIA GAM (Diagnostica Stago), and Platelet lysate-Heparin-Zymutest HIA IgG immunoassay (Hyphen BioMed)] are shown. Additionally, PF4-dependent P-selectin activation assay (PEA), and serotonin release assay (SRA) functional testing results are provided. All results are correlated with each patient’s clinically defined probability of HIT (4Ts score). Positive test results are denoted in red font, negative results in black font. Positive assay cut-offs vary by run for the Asserachrom HPIA GAM and are enclosed in parentheses. Positive cut-offs for Lifecodes PF4 IgG, Lifecodes PF4 GAM, Zymutest HIA IgG, SRA and PEA are 0.4 OD, 0.4 OD, 0.3 OD, 20% and 19%, respectively.

Prior approaches to develop murine-derived HIT monoclonal antibodies have solely relied on PF4/polyanion ELISA-based screens, which, by design, would not lead to the development of ELISA-negative platelet-activating anti-PF4 antibodies^17^. Here, a novel dual-screening strategy was employed, whereby antibodies were simultaneously screened in a functional assay (PEA) and an antigenic assay (PF4/polyanion ELISA), to identify PF4-dependent platelet-activating antibodies that do not recognize PF4-polyanion complexes. One ELISA-negative monoclonal antibody (EN-mAb) was identified that had serologic characteristics indistinguishable from pathogenic ELISA-negative HIT patient antibodies: EN-mAb did not bind PF4-polyanion targets, both PF4/PVS and PF4/heparin, in HIT immunoassays (**Fig. 4A**). In contrast, the well-characterized HIT-like murine mAb KKO, bound to both PF4/PVS and PF4/heparin complexes (**Fig. 4A**). In functional assays employing PF4-treated platelets, both EN-mAb and KKO stimulated robust PF4-dependent platelet activation, which was inhibited by high concentrations of heparin, characteristic of HIT antibodies (**Fig. 4B**). Consistent with platelet activation results, both EN-mAb and KKO bound platelets in a PF4-dependent manner (**Fig. 4C**).

**Figure 4.**
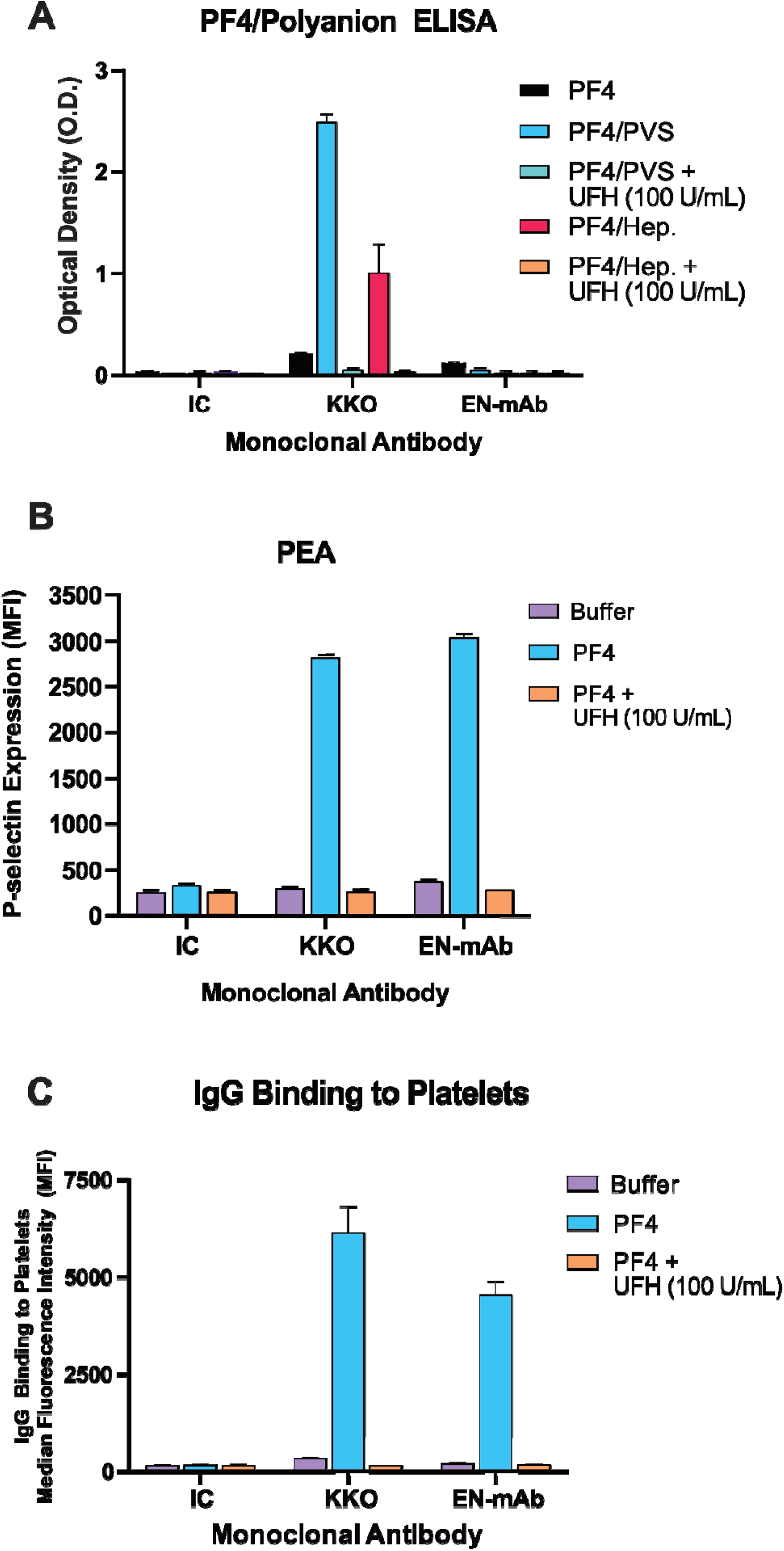
Serologic characterization of an ELISA-negative, platelet-activating murine monoclonal antibody. Serological characterization of the murine monoclonal antibody (EN-mAb) was performed using HIT ELISA, platelet activation assays, and IgG-platelet binding studies. **(A)** HIT immunoassays were performed to assess the binding of the well-characterized “HIT-like” murine monoclonal antibody (KKO), EN-mAb, or IC (isotype control murine monoclonal antibody) to several different immobilized antigens: PF4, PF4/polyvinylsulfonate (PVS) polyanion complexes, PF4/PVS polyanion complexes with the addition of high concentration (100 U/mL) of unfractionated heparin (UFH), PF4/heparin, and PF4/heparin with high concentration of UFH. **(B)** Platelet activation (P-selectin expression) induced by KKO, EN-mAb, and IC was evaluated in platelets treated with buffer, PF4, or PF4 and excess (100 U/mL) UFH. **(C)** Antibody binding to platelets was quantified for EN-mAb, KKO, and IC using platelets incubated with buffer, PF4, or PF4-treated platelets with excess (100 U/mL) unfractionated heparin (UFH). The mean of triplicate values with error bars representing one standard deviation is displayed for all studies.

## DISCUSSION

Our findings of ELISA-negative pathogenic PF4-dependent platelet-activating antibodies in a recently identified anti-PF4 antibody-mediated disorder, MGTS^18^, led us to explore antibodies with this unusual serology among HIT-suspected patients. We initially identified three cases of HIT that were negative in one of the most commonly performed HIT ELISA assays (Lifecodes PF4 IgG, Immucor)^12^, which uses PF4/polyvinylsulfonate (PVS) targets for HIT antibody detection. Notably, in addition to ELISA diagnostic kits, both automated HIT tests approved for diagnostic use in the United States, the LIA and CLIA, also utilize PF4/PVS complexes as targets (sample volume remaining were inadequate for testing in these additional assays)^19^. All patients had clinical histories highly consistent with HIT, and functional testing confirmed the presence of platelet-activating PF4-specific antibodies, and the data clearly demonstrated PF4-dependent binding of these antibodies to platelets. To further investigate the incidence of ELISA-negative HIT patients with platelet-activating anti-PF4 antibodies, an unbiased retrospective screen of 500 consecutive PF4/PVS ELISA-negative samples from HIT-suspected patients was undertaken, which yielded an incidence of 0.6% of ELISA-negative platelet-activating HIT antibodies (3/500 patients). Additional studies are needed to verify the prevalence of this class of antibodies.

Additional testing for IgA and IgM antibodies to PF4/PVS complexes yielded weak positive reactions (OD < 1.0) in two of the six patients. The pathogenicity of IgA and IgM antibodies in HIT remains unclear, as platelets lack receptors for these antibody types. Testing for IgG, IgA, and IgM antibodies against PF4/heparin targets in the only FDA-approved PF4/heparin ELISA identified two of the six patient antibodies (one patient had an OD <1.0). Experts recommend the use of an OD cut-off of 1.0 for a positive test^20^, thus, only one patient was deemed likely to have a pathogenic antibody using this test. Interestingly, five of the six samples harbored antibodies to heparin-platelet lysate mixtures, suggesting that the presence of cofactors in platelet lysate may facilitate the recognition of PF4 by HIT antibodies, or indicating the presence of antibodies to other platelet antigens (e.g., IL-8^21,22^, NAP-2). However, apart from EN-HIT3, where antibodies to IL-8 were demonstrated, no antibodies to IL-8 were observed in any other patient, and no antibodies to NAP-2 were detected.

To our knowledge, there are only two single-case reports of a bona fide pathogenic HIT ELISA-negative platelet-activating HIT antibody in the published literature^23,24^. In the first study^23^, the authors investigated the phenomenon of platelet activation-positive/ELISA-negative HIT in a large cohort of patients (>8000) and, other than the index case, found no instances of additional pathogenic ELISA-negative HIT patients, suggesting that this entity was “exceptionally uncommon”. In the second study, the patient presented with a clinical picture highly consistent with HIT with positive SRA serology, despite being ELISA-negative^24^, and no additional cases were evaluated. In a third study of more than 1,500 patients^25^, two patients with platelet activation-positive/ELISA-negative cases were identified. However, the authors concluded that reasons other than HIT explained the isolated thrombocytopenia seen in these two cases, suggesting that these antibodies were non-pathogenic. Lastly, in a study of >1000 patients, ELISA-negative SRA-positive samples were identified in 0.7% of patients^26^, a finding similar to the estimates in this report. However, no clinical information was provided to adjudicate whether the patients had a clinical course consistent with HIT. Our results contrast with prior studies^23,25^ in that we found a higher prevalence of platelet-activating, ELISA-negative antibodies. One potential explanation for this difference is that the functional screen performed in this study utilized platelets treated with PF4. In contrast, prior studies performed SRA/HIPA testing, which use heparin-treated platelets. In fact, one of the patients in our study (EN-HIT4) was deemed to have “SRA-negative HIT” with activating antibodies identified only through PF4-based functional testing. This entity is now well recognized^27,28^.

A key objective to better understand the PF4 epitopes recognized by this class of antibodies was to generate monoclonal antibodies that recapitulate the platelet-activating but ELISA-negative serologic profile of patient antibodies. To this end, we successfully developed a PF4-dependent platelet-activating antibody, which does not recognize PF4/polyanion complexes using a novel ELISA and functional testing dual-screening strategy.

Current practice guidelines, such as those from the American Society of Hematology^11^, recommend discontinuation of non-heparin anticoagulation and resumption of heparin, if indicated, in patients with intermediate-probability 4Ts scores (recommendation 2.7) or high-probability 4Ts scores (recommendation 2.8) and a negative immunoassay. Two of the six HIT patients in our cohort had intermediate-probability 4Ts scores and, in line with recommendation 2.7, were not subject to significant additional testing before heparin reintroduction, which resulted in thrombocytopenia. Despite a qualifying “remark” in recommendation 2.8, which suggests a different immunoassay, testing in a functional assay, and evaluation for antibodies to non-PF4 antigens (e.g., IL-8, NAP-2), three of the four high 4Ts scored ELISA-negative HIT patients were re-exposed to heparin after the interruption of direct thrombin inhibitors, resulting in a decrease in platelet count in all patients and the development of new thrombosis in one (EN-HIT1). Non-implementation of this guidance likely reflects the real-world challenges in accessing a different PF4/polyanion immunoassay (e.g., typically a hospital lab only has one validated frontline HIT assay), availability of functional testing (usually only available in reference laboratories), and lack of availability of standardized tests for IL-8 and NAP-2 antibodies. Failing to recognize ELISA-negative, platelet-activating HIT antibodies risks patient harm by triggering cessation of non-heparin anticoagulation treatments and re-exposure to heparin. Thus, a careful reevaluation of practice guidelines may be warranted to strike a balance between recognizing ELISA-negative HIT and the risk of over-testing for HIT.

## Data Availability

All data produced in the present work are contained in the manuscript and supplemental file.

## Acknowledgments

We thank Stephanie Hafner and Jill Kappers from Mayo Clinic’s Research Innovation Office for their exceptional support in research coordination. This work was supported, in part, by National Institutes of Health grants HL158932 (AP), HL171911 (AJK), HL147734 (CGJ), HL148120 (RW), HL167668 (RW, DW) and HL130724 (DW).

## Authorship

AJK, NPS, EEM, and RRL performed the laboratory studies. LT, MG, SJ, AG, KJS, HA, AA, NLR, and AIC provided critical clinical correlates and helpful feedback. RHA provided helpful advice on the development of murine monoclonal antibodies. EY, CGJ, and SMP collaborated on the development of murine antibodies. DW and RW oversaw NAP-2 and IL-8 testing. MCK, RRL, DLM, DC, DW, RW, and RKP provided valuable feedback regarding the clinical and laboratory aspects of the manuscript. AJK and AP conceived the experimental plan and directed the laboratory studies. AJK and AP wrote the first draft; all authors provided input and approved the final version.

## REFERENCES

1. Arepally GM, Padmanabhan A. Heparin-Induced Thrombocytopenia: A Focus on Thrombosis. Arterioscler Thromb Vasc Biol 2021;41(1):141–152. DOI: 10.1161/ATVBAHA.120.315445.

2. Cuker A. Clinical and laboratory diagnosis of heparin-induced thrombocytopenia: an integrated approach. Semin Thromb Hemost 2014;40(1):106–14. (In eng). DOI: 10.1055/s-0033-1363461.

3. Warkentin TE. How I diagnose and manage HIT. Hematology Am Soc Hematol Educ Program 2011;2011:143–9. (In eng). DOI: 10.1182/asheducation-2011.1.143.

4. Arepally GM, Ortel TL. Vaccine-induced immune thrombotic thrombocytopenia: what we know and do not know. Blood 2021;138(4):293–298. DOI: 10.1182/blood.2021012152.

5. Greinacher A, Selleng K, Palankar R, et al. Insights in ChAdOx1 nCoV-19 vaccine-induced immune thrombotic thrombocytopenia. Blood 2021;138(22):2256–2268. DOI: 10.1182/blood.2021013231.

6. Huynh A, Kelton JG, Arnold DM, Daka M, Nazy I. Antibody epitopes in vaccine-induced immune thrombotic thrombocytopaenia. Nature 2021;596(7873):565–569. DOI: 10.1038/s41586-021-03744-4.

7. Vayne C, Rollin J, Gruel Y, et al. PF4 Immunoassays in Vaccine-Induced Thrombotic Thrombocytopenia. N Engl J Med 2021;385(4):376–378. DOI: 10.1056/NEJMc2106383.

8. Kanack AJ, Singh B, George G, et al. Persistence of Ad26.COV2.S-associated vaccine-induced immune thrombotic thrombocytopenia (VITT) and specific detection of VITT antibodies. Am J Hematol 2022;97(5):519–526. DOI: 10.1002/ajh.26488.

9. Kanack AJ, Bayas A, George G, et al. Monoclonal and oligoclonal anti-platelet factor 4 antibodies mediate VITT. Blood 2022;140(1):73–77. DOI: 10.1182/blood.2021014588.

10. Kanack AJ, Schaefer JK, Sridharan M, et al. Monoclonal gammopathy of thrombotic/thrombocytopenic significance. Blood 2023;141(14):1772–1776. DOI: 10.1182/blood.2022018797.

11. Cuker A, Arepally GM, Chong BH, et al. American Society of Hematology 2018 guidelines for management of venous thromboembolism: heparin-induced thrombocytopenia. Blood Adv 2018;2(22):3360–3392. DOI: 10.1182/bloodadvances.2018024489.

12. Liederman Z, Van Cott EM, Smock K, Meijer P, Selby R. Heparin-induced thrombocytopenia: An international assessment of the quality of laboratory testing. J Thromb Haemost 2019;17(12):2123–2130. DOI: 10.1111/jth.14611.

13. Samuelson Bannow B, Warad DM, Jones CG, et al. A prospective, blinded study of a PF4-dependent assay for HIT diagnosis. Blood 2021;137(8):1082–1089. DOI: 10.1182/blood.2020008195.

14. Padmanabhan A, Jones CG, Curtis BR, et al. A Novel PF4-Dependent Platelet Activation Assay Identifies Patients Likely to Have Heparin-Induced Thrombocytopenia/Thrombosis. Chest 2016;150(3):506–15. DOI: 10.1016/j.chest.2016.02.641.

15. Padmanabhan A, Jones CG, Bougie DW, et al. Heparin-independent, PF4-dependent binding of HIT antibodies to platelets: implications for HIT pathogenesis. Blood 2015;125(1):155–61. DOI: 10.1182/blood-2014-06-580894.

16. Kanack AJ, Jones CG, Singh B, et al. Off-the-shelf cryopreserved platelets for the detection of HIT and VITT antibodies. Blood 2022;140(25):2722–2729. DOI: 10.1182/blood.2022017283.

17. Arepally GM, Kamei S, Park KS, et al. Characterization of a murine monoclonal antibody that mimics heparin-induced thrombocytopenia antibodies. Blood 2000;95(5):1533–1540. (In English). DOI: DOI 10.1182/blood.V95.5.1533.005k01_1533_1540.

18. Kanack AJ, Leung N, Padmanabhan A. Diagnostic Complexity in Monoclonal Gammopathy of Thrombotic Significance. N Engl J Med 2024;391(20):1961–1963. DOI: 10.1056/NEJMc2409428.

19. Warkentin TE. Laboratory diagnosis of heparin-induced thrombocytopenia. Int J Lab Hematol 2019;41 Suppl 1:15–25. DOI: 10.1111/ijlh.12993.

20. Chan CM, Woods CJ, Warkentin TE, Sheppard JI, Shorr AF. The Role for Optical Density in Heparin-Induced Thrombocytopenia: A Cohort Study. Chest 2015;148(1):55–61. (In eng). DOI: 10.1378/chest.14-1417.

21. Regnault V, de Maistre E, Carteaux JP, et al. Platelet activation induced by human antibodies to interleukin-8. Blood 2003;101(4):1419–21. DOI: 10.1182/blood-2002-02-0620.

22. Amiral J, Marfaing-Koka A, Wolf M, et al. Presence of autoantibodies to interleukin-8 or neutrophil-activating peptide-2 in patients with heparin-associated thrombocytopenia. Blood 1996;88(2):410–6. (https://www.ncbi.nlm.nih.gov/pubmed/8695787).

23. Warkentin TE, Smythe MA, Ali MA, et al. Serotonin-release assay-positive but platelet factor 4-dependent enzyme-immunoassay negative: HIT or not HIT? Am J Hematol 2021;96(3):320–329. DOI: 10.1002/ajh.26075.

24. Attah A, Peterson C, Jacobs M, et al. Anti-PF4 ELISA-Negative, SRA-Positive Heparin-Induced Thrombocytopenia. Hematol Rep 2024;16(2):295–298. DOI: 10.3390/hematolrep16020029.

25. Greinacher A, Juhl D, Strobel U, et al. Heparin-induced thrombocytopenia: a prospective study on the incidence, platelet-activating capacity and clinical significance of antiplatelet factor 4/heparin antibodies of the IgG, IgM, and IgA classes. J Thromb Haemost 2007;5(8):1666–73. (In eng). DOI: 10.1111/j.1538-7836.2007.02617.x.

26. Shelat SG, Tomaski A, Pollak ES. Serologic results in >1000 patients with suspected heparin-induced thrombocytopenia. Clin Appl Thromb Hemost 2008;14(4):410–4. DOI: 10.1177/1076029607304721.

27. Pandya KA, Johnson EG, Davis GA, Padmanabhan A. Serotonin release assay (SRA)-negative HIT, a newly recognized entity: Implications for diagnosis and management. Thromb Res 2018;172:169–171. DOI: 10.1016/j.thromres.2018.10.022.

28. Warkentin TE, Nazy I, Sheppard JI, Smith JW, Kelton JG, Arnold DM. Serotonin-release assay-negative heparin-induced thrombocytopenia. Am J Hematol 2020;95(1):38–47. DOI: 10.1002/ajh.25660.

