## Supplementary Appendix for "Pathogenic PF4/Polyvinylsulfonate ELISA-negative Antibodies in HIT"

**Table of Contents**

**Methods…………………………………………………………………………………………………1**

**Detailed Patient Histories……………………………………………………………………………4**

**Figures…………………………………………………………………………………………………..7**

**References…………………………………………………...…………………………………………9**

***METHODS***

***Functional studies***

The PF4-dependent P-selectin expression assay was performed as previously described^1^. Prostaglandin E1 was added to citrated whole blood from healthy volunteers at a final concentration of 50 ng/mL, and the whole blood was centrifuged at 200 x *g* for 15 minutes to obtain platelet-rich plasma (PRP). Platelets were then isolated from PRP by centrifugation at 1,000 x *g* for 10 minutes by resuspending the platelet pellet in phosphate-buffered isotonic saline (pH 7.4) supplemented with 1% bovine serum albumin (PBS-BSA). Platelets were then incubated with ten microliters of patient sample for one hour at ambient temperature before adding fluorescently labeled anti-P-selectin (monoclonal antibody HB-299, ATCC) and anti-GPIIIa (monoclonal antibody HB-242, ATCC) antibodies for 20 minutes. PBS-BSA was then added to a final volume of 200 µL. Platelet events were collected after gating based on GPIIIa positivity. P-selectin expression (median fluorescence intensity, MFI) was measured as a marker of platelet activation. In some studies, platelets were incubated with heparin (100 U/mL), the anti-FcγRIIa receptor monoclonal antibody IV.3, or an isotype control monoclonal antibody at a concentration of two µg/mL prior to PEA testing. Testing in the Thrombospondin-1 release assay using cryopreserved platelets was performed as previously described^2^, with minor modifications.

***IgG-Platelet Binding Studies***

For patient samples, platelet binding assays were performed as described previously^3^. Prostaglandin E1 (50 ng/mL) was added to healthy donor whole blood, and PRP was isolated by centrifugation at 200 × g for 15 minutes. PRP was centrifuged at 1000 × g for 10 minutes to pellet platelets before platelet resuspension in PBS-BSA. Next, 1.0 × 10^6^ platelets were treated with serum samples (10 µL) to produce a final reaction mixture of 50 µL. After incubation for 15 minutes at ambient temperature, platelets were centrifuged once at 1000 × g and then resuspended in 50 µL PBS-BSA containing APC-labeled goat anti-human IgG (1:100). The mixture was then incubated in the dark for 30 minutes, samples were diluted to 250 μL, and fluorescence was quantified. For murine-derived monoclonal antibodies, the antibodies were directly labeled with AlexaFluor 647 according to manufacturer instructions (Invitrogen), and binding of the antibodies to platelets was assessed using untreated platelets or platelets treated for 20 minutes with 30 µg/mL PF4 or 30 µg/mL PF4 with 100U/mL heparin for 20 minutes. Samples were analyzed using an Accuri C6 flow cytometer.

***ELISA studies***

ELISA plates (Thermo Scientific) were incubated with recombinant PF4 and polyvinyl sulfonate (PVS, Polysciences; 9 µg/mL) or PF4 and unfractionated heparin (0.4 U/mL). Plates were washed with PBS with 0.1% Tween-20 and blocked with Superblock T20 (Thermo Scientific). Samples were tested at a 1:50 dilution, and murine monoclonals at four µg/mL. AP-labelled goat anti-human IgG Fc or goat anti-mouse IgG Fc antibodies (Jackson Immunoresearch; 1:5000) and pNPP (Sigma Aldrich) were used for colorimetric detection. Optical density was recorded at 30 minutes at 405 and 492 nanometers. In some studies, serum samples were incubated with high concentrations of heparin (100 U/mL).

The binding of patient IgG to NAP-2 or IL-8 was assessed by ELISA using methods adapted from previously published protocols^4,5^. Briefly, 96-well microwells were coated overnight at ambient temperature with 50 μL of one μg/mL IL-8 (Protein Foundry) or NAP-2 (provided by Dr. Mortimer Poncz laboratory) after dilution in PBS. After washing the plates with PBS and 0.1% Tween-20, each well of interest was blocked for two hours at ambient temperature using PBS supplemented with 3% BSA and 0.1% Tween-20 (PBS-BSA). Patient samples diluted 1:100 in PBS-BSA (50 μL per well) were added and incubated for one hour at ambient temperature with orbital shaking. Plates were washed, followed by the addition of 50 μL HRP-conjugated goat anti-human IgG in dilution buffer (200 ng/mL, Southern Biotech) and incubation for 45 min at ambient temperature. After a final wash, TMBZ substrate (A+B, KPL, Sera Care) was added, and color development was allowed for 15 minutes at ambient temperature in the dark. The reaction was stopped with 50 μL of 1 N H₂SO₄, and absorbance at 450 nm was measured to quantify antibody binding to immobilized proteins.

***Development of murine-derived monoclonal antibody***

Three mice were immunized with 50 µg of PF4 and 3U of heparin at 0, 4, and 6 weeks, then once every four weeks. A dual screening approach employing both PF4/polyvinyl sulfonate (PVS) immunoassay and PEA testing was then used to select hybridoma clones of interest, including those that were PF4/PVS immunoassay-negative but PEA-positive. After selecting clones, mAbs were purified using protein G Sepharose (Cytiva Life Sciences) according to manufacturer’s instructions. The purified anti-PF4, isotype control, or KKO antibodies at two µg/mL were then tested against PF4/PVS, or PF4/PVS targets with 100 U/mL heparin in ELISA and in functional studies as described above.

***DETAILED PATIENT HISTORIES***

***Clinical presentation and course of patients identified by functional screening.***

The first ELISA-negative HIT cases identified in the screen (EN-HIT4; **Fig. 2C**) was a female in her 50s who received a liver transplant after a diagnosis of hilar cholangiocarcinoma and was treated with low molecular weight heparin (LMWH) on post-operative days 1-5 and then low-dose aspirin on post-operative days 10-12 following the surgical procedure. The patient developed significant thrombocytopenia ten days post-transplant, which subsequently recovered over the next ~10 days. She was subsequently noted to have extensive portal vein thrombosis. She was started on a heparin drip, during which the patient’s platelet counts rapidly decreased to a platelet nadir of 42x10^3^/µL. The 4Ts score was 7. At this time, a HIT ELISA was performed and returned a negative result (OD, 0.112); however, functional testing was not performed. Retrospectively, the patient’s residual stored sample was tested and returned negative in the SRA (<5%), despite a positive result in PEA (**Figs. 2A** & **2B**). Over the next several weeks, the patient's platelet count normalized with a transition to a Factor Xa inhibitor. However, the patient was re-admitted to the hospital 52 days post-transplant for volume overload. Following admission, imaging revealed progression of portal vein thrombosis, and the patient was again anticoagulated using LMWH and UFH, which resulted in another significant decrease in platelet counts (**Fig. 2C**). The cessation of heparin treatment was associated with a robust rebound in the platelet count, further supporting a diagnosis of HIT.

EN-HIT5 (**Fig. 2D**) was a female in her 60s who received heparin during cardiac surgery for aortic and mitral tissue valve replacements. Following surgery, the patient was given three doses of subcutaneous heparin as DVT prophylaxis. Once bleeding was noted to have stopped, bivalirudin (per surgeon preference) was initiated as a bridge to systemic anticoagulation with warfarin. Bivalirudin was maintained at therapeutic levels via activated partial thromboplastin time (APTT) assessments. Anticoagulation management was complicated by one instance of a supratherapeutic INR, for which bivalirudin/warfarin were temporarily interrupted (**Fig. 2D**). Treatment with heparin was followed by a significant decrease in platelet counts. Due to thrombocytopenia and a history of recent treatment with heparin, HIT ELISA testing was performed and came back with a negative result (0.168 OD). The 4Ts score was 5. Platelet counts steadily recovered until additional heparin exposures occurred 34 and 45 days post-surgery, at which point the patient experienced significant decreases in platelet counts coinciding with heparin reexposures, but with no known thrombotic events. SRA testing performed retrospectively on the sample that tested ELISA-negative was strongly positive at 90%.

EN-HIT6 (**Fig. 2E**) was a male in his 70s diagnosed with metastatic renal cell carcinoma who was admitted due to acute visual loss with homonymous hemianopsia due to multiple cerebral infarcts. The patient was treated acutely with tenecteplase, and his course was complicated by hemorrhagic transformation. He was found to have a bidirectional patent foramen ovale but no deep vein thrombosis in the lower extremities. He was started on heparin, and after a brief interruption for posterior fusion surgery of vertebrae T4-L2 and resection of a cancerous mass on T11, he was continued on unfractionated heparin as prophylaxis for venous thromboembolism. A steady decline of platelet counts was noted during and following his treatment with heparin. During this period, the patient was found to have a submassive pulmonary embolism with right ventricular dysfunction by echocardiography, extensive thrombosis of the right lower extremity extending from the calf veins into the common femoral vein, as well as right renal and acute myocardial infarct (both attributed to paradoxical embolization in the setting of bidirectional cardiac shunt). At the time of these events, the patient was on prophylactic unfractionated heparin and experienced a platelet drop from 233x10^3^/µL to 152x10^3^/µL and a further decline to 96x10^3^/µL within 12 hours of starting IV heparin to treat the acute thrombotic events. Heparin was discontinued in favor of a DTI while HIT ELISA testing was pending. The 4Ts score was calculated at 5. Upon confirmation of HIT ELISA negativity (OD, 0.159), the DTI was discontinued, and the patient was transitioned briefly to LMWH, which resulted in a decrease in platelet count. After a period of treatment with a Factor Xa inhibitor, which resulted in significant platelet recovery (to 124 x 10^3^/µL), brief re-exposure to heparin after a second ELISA tested negative (0.141 OD) corresponded with a platelet decrease from 124 to 79 x 10^3^/µL after which the patient was transitioned back to a Factor Xa inhibitor. Shortly after transitioning to an FXa inhibitor, platelets increased rapidly and were at 201x10^3^/µL seventy-two hours later.

**F**
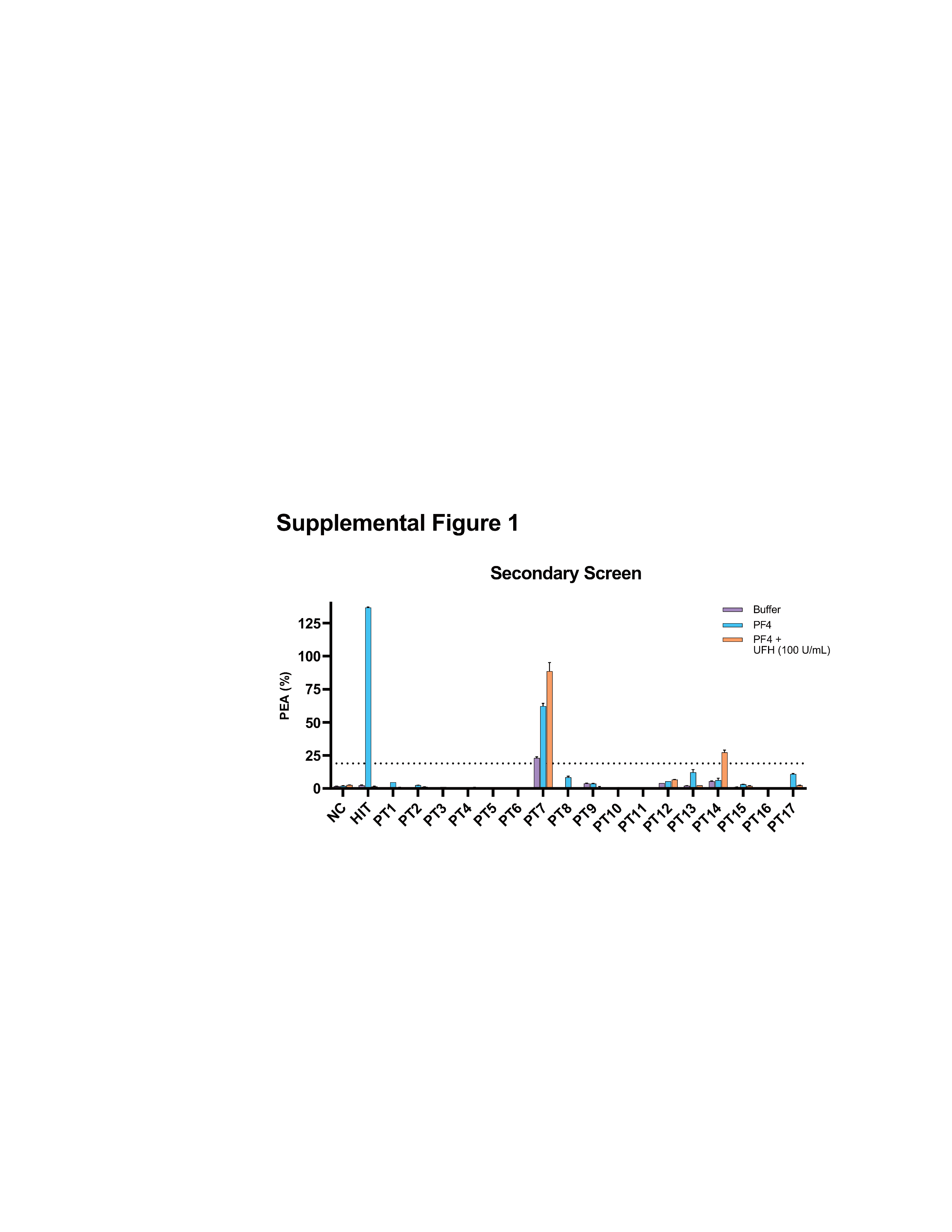
**IGURES**

**Figure S1. Secondary testing of HIT-suspected patients in a PF4-dependent functional assay.** An initial screen of 500 consecutive ELISA-negative HIT-suspected patients yielded 20 samples that stimulated platelet activation (≥19%) in the PF4-dependent P-selectin expression assay (PEA; **Fig. 2A**). To assess PF4-dependent and high-concentration heparin-inhibitable reactions that would be characteristic of HIT antibodies, secondary testing was performed using platelets from a second, independent platelet donor under several conditions: buffer-treated platelets, PF4-treated platelets, and PF4-treated platelets with 100 U/mL unfractionated heparin (UFH). Seventeen patients (PT1-17) failed to stimulate PF4-dependent platelet activation, which was inhibited by high heparin concentrations. NC- negative control. HIT- HIT positive control.

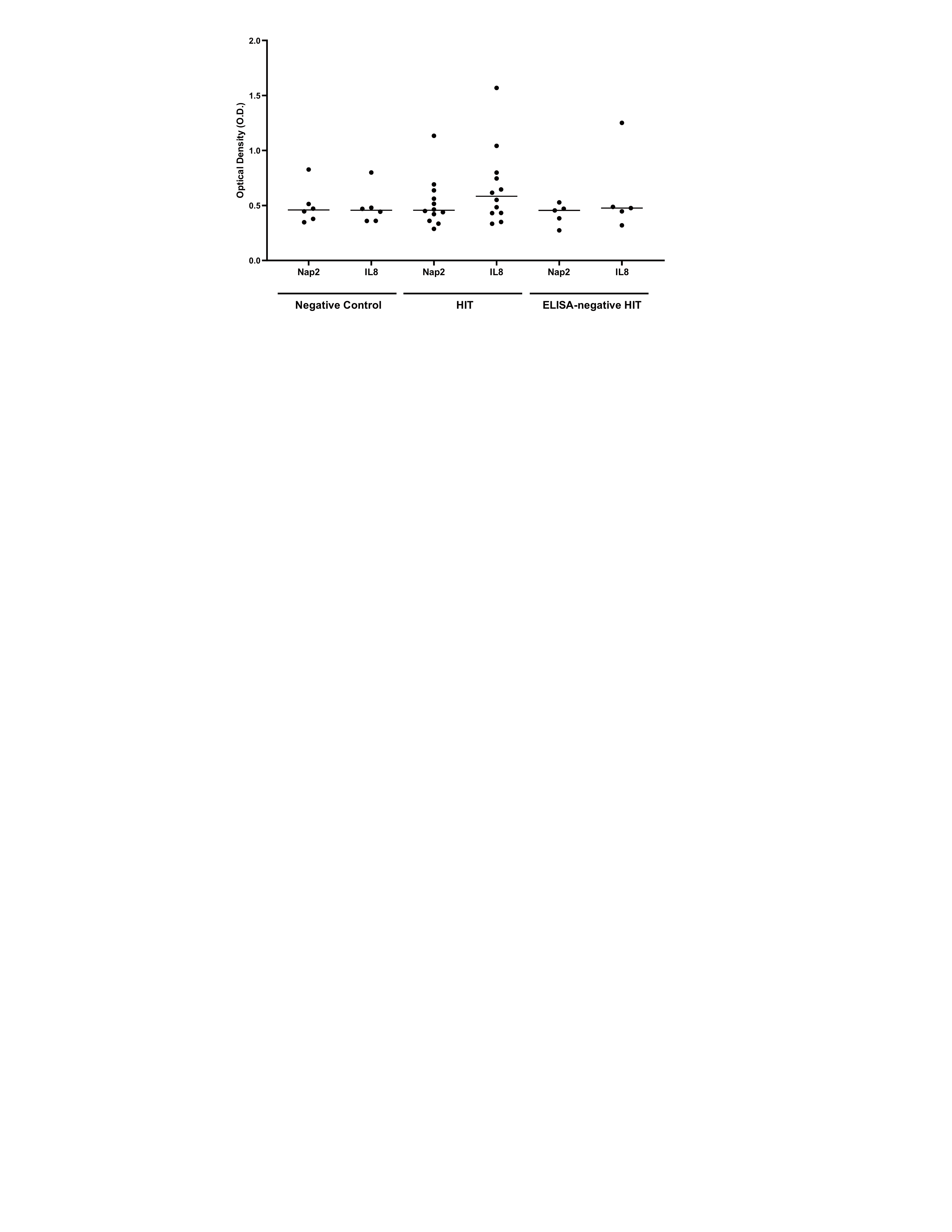

**Figure S2. ELISA assay to measure immunoglobulin G binding to NAP-2 and IL-8.** Samples from healthy donors (negative controls), confirmed HIT patients, or ELISA-negative HIT patients (EN-HIT 1 through 5) were assessed for IgG binding to recombinant IL-8 or Nap-2.

**References**

1.	Samuelson Bannow B, Warad DM, Jones CG, et al. A prospective, blinded study of a PF4-dependent assay for HIT diagnosis. Blood 2021;137(8):1082-1089. DOI: 10.1182/blood.2020008195.

2.	Kanack AJ, Jones CG, Singh B, et al. Off-the-shelf cryopreserved platelets for the detection of HIT and VITT antibodies. Blood 2022;140(25):2722-2729. DOI: 10.1182/blood.2022017283.

3.	Padmanabhan A, Jones CG, Bougie DW, et al. Heparin-independent, PF4-dependent binding of HIT antibodies to platelets: implications for HIT pathogenesis. Blood 2015;125(1):155-61. DOI: 10.1182/blood-2014-06-580894.

4.	Zhu W, Zheng Y, Yu M, et al. Cloned antibodies from patients with HIT provide new clues to HIT pathogenesis. Blood 2023;141(9):1060-1069. DOI: 10.1182/blood.2022017612.

5.	Zhu W, Zheng Y, Yu M, et al. Prothrombotic antibodies targeting the spike protein's receptor-binding domain in severe COVID-19. Blood 2025;145(6):635-647. DOI: 10.1182/blood.2024025010.
